# The Impact of a Primary Aldosteronism Predictive Model in Secondary Hypertension Decision Support

**DOI:** 10.1101/2024.07.09.24310088

**Authors:** Peter B. Mack, Casey Cole, Mintaek Lee, Lisa Peterson, Matthew Lundy, Karen Hegarty, William Espinoza

## Abstract

**Objective:** To determine whether the addition of a primary aldosteronism (PA) predictive model to a secondary hypertension decision support tool increases screening for PA in a primary care setting.

**Materials and Methods:** 153 primary care clinics were randomized to receive a secondary hypertension decision support tool with or without an integrated predictive model between August 2023 and April 2024.

**Results:** For patients with risk scores in the top 1 percentile, 63/2,896 (2.2%) patients where the alert was displayed in model clinics had the order set launched while 12/1,210 (1.0%) in no model clinics had the order set launched (P = 0.014). 19/2,896 (0.66%) of these highest risk patients in model clinics had an ARR ordered compared to 0/1,210 (0.0%) patients in no model clinics (P = 0.010). For patients with scores not in the top 1 percentile, 438/20,493 (2.1%) patients in model clinics had the order set launched compared to 273/17,820 (1.5%) in no model clinics (P < 0.001). 124/20,493 (0.61%) in model clinics had an ARR ordered compared to 34/17,820 (0.19%) in the no model clinics (P < 0.001).

**Discussion:** The addition of a PA predictive model to secondary hypertension alert displays and triggering criteria along with order set displays and order preselection criteria results in a statistically and clinically significant increase in screening for PA, a condition that clinicians insufficiently screen for currently.

**Conclusion:** Addition of a predictive model for an under-screened condition to traditional clinical decision support may increase screening for these conditions.

## 1. Introduction

Primary aldosteronism (PA) is a treatable cause of secondary hypertension that requires targeted screening tests to detect; tests that are performed with suboptimal frequency. Current hypertension guidelines recommend health care providers consider secondary hypertension in all patients with a new diagnosis of hypertension. PA is thought to be present in 5-10% of patients with hypertension, although there is considerable uncertainty as to the true prevalence.^1-5^

It is challenging to determine how many patients with hypertension have been screened for all forms of secondary hypertension as discrete documentation about the consideration of causes like medication-induced hypertension and alcohol-induced hypertension are rarely present in the medical record. There is, however, clear data that PA is screened for in only a small fraction of patients even with resistant hypertension.^6,7^ Identifying patients with PA is important as performing an adrenalectomy on patients with PA has significant therapeutic advantages over medical treatment^8^ and surgical treatment would not even be considered without an appropriate diagnosis.

Screening for PA is performed by measuring an aldosterone to plasma renin activity ratio (ARR). Many commonly prescribed antihypertensives will impact the measurement of the ARR, but only a small number of antihypertensives are contraindicated when measuring the ARR.^9^ Treatment of confirmed PA consists of either a unilateral adrenalectomy or a mineralocorticoid antagonist depending on laterality of the disease and whether the patient is amenable to a surgical approach.^10^ Given the complex diagnosis and management of PA, clinical decision support tools have the potential to improve quality of care, particularly in a primary care setting where most hypertension is managed,^11^ but where physicians face intense time pressure^12^ which hinders their ability to perform more complex screening tests.

The value of non-interruptive alerts to help remind health care providers to perform screening for PA via order sets has been demonstrated.^13^ Predictive risk models have been published to help identify patients at risk of having PA,^7,14^ to make a diagnosis of PA when screening data is available,^15^ to identify patients who have unilateral PA,^16-18^ and to predict resolution of hypertension after adrenalectomy in PA.^19-21^ With the proliferation of risk models related to PA, the next important problem to solve becomes the communication of the risk model results and the effective integration of the risk model into the clinical workflow. A risk model was integrated with a comprehensive secondary hypertension clinical decision support system to determine incremental value of adding a risk model to decision support.

### 2. Methods

### 2.1. PA predictive model

A logistic regression model^22^ was created to predict the probability of a patient having an increased ARR (value > 35) resulting in 10% positive rate. A total of 27 features including age, sex, problem-list diagnoses, prescriptions, lab parameters, and blood pressure were considered for the model. These features were selected for consideration based on other models for aldosteronism subtype classification^16-18^ and known factors important in hypertension management and subtype classification. Data was extracted from 3,746 patients and missing data was imputed by K-Nearest Neighbors where k=4 and neighbor weight is determined by distance.^23^ From these 27 candidate features, 18 features were selected using Hilbert-Schmidt independence criterion least absolute shrinkage and selection operator (HSIC LASSO)^24^ that best independently discriminated between increased and normal ARR values. The model had an AUROC of 0.77, an AUPRC of 0.3, and an expected calibration error^25^ of 0.03. Fairlearn analysis was utilized to ensure that there was little to no prediction bias across sensitive patient attributes. The model was found to produce equitable predictions across sex, race, and age groups.

### 2.2. Alert design

A non-interruptive was built for potential secondary hypertension in Novant Health’s EHR triggered based on the quantitative elements of the 2017 ACC hypertension guidelines along with predictive model scores in the top 1 percentile (Figure 1). The selected triggers from the guideline were 1) age less than 30 with a blood pressure above 140/90 or a diagnosis of hypertension, 2) a blood pressure above 180/110, 3) a diastolic blood pressure above 90 mmHg in a patient aged 65-years-old or older, 4) both a current and recent average blood pressure above 140/90 despite being on three or more different classes of antihypertensive agents. The alert displays the reason or reasons why it was triggered to display and in patients with predictive model scores in the top 1 percentile displays, “Patient may have a greatly increased risk of an abnormal aldosteronism screening test based on a predictive model” (also a trigger) while for patients in the top 5 percentile, but not the top 1 percentile displayed, “Patient may have an increased risk of an abnormal aldosteronism screening test based on a predictive model” (not a trigger). This alert includes a link to a dynamic secondary hypertension order set.

**Figure 1.**
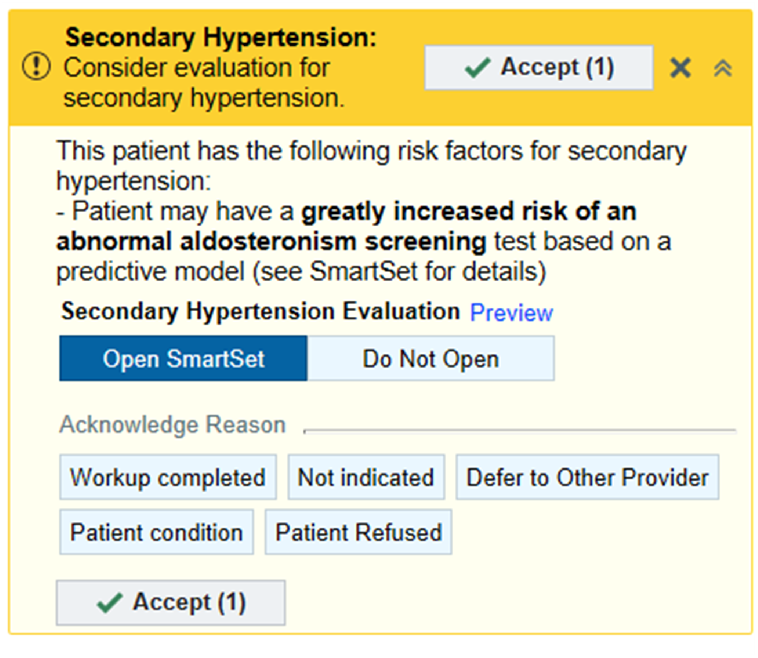
Non-interruptive alertexample based on synthetic data.

### 2.3. Order set design

The order set contains two main sections, a summary of key recommendations specific to the patient and evaluation tools for individual causes of secondary hypertension (Figure 2). The key recommendations section contains the aforementioned text from the alert denoting whether a patient had an “increased” or “greatly increased” risk of abnormal aldosteronism screening but does not contain any additional text when the risk score was below the 95th percentile. When any increased risk was denoted, the three most significant factors contributing to this risk estimate were listed. Other secondary hypertension recommendations that conditionally display include guidance for suspected alcohol-induced hypertension,^26^ unevaluated hypercalcemia, untreated hyperparathyroidism,^27^ an unevaluated elevated ARR, or an ARR suggestive of pseudoaldosteronism.

**Figure 2.**
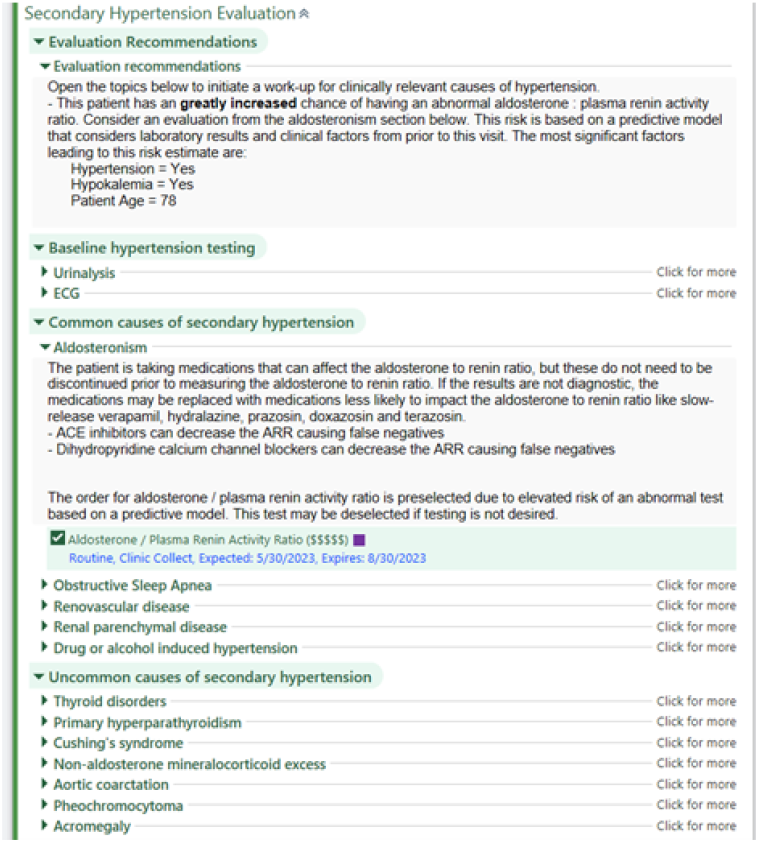
Order set example based on synthetic data.

The remainder of the order set contains the major categories of secondary hypertension from the ACC guidelines.^1^ When expanded, each section contains brief explanatory text as well as any necessary orders. The “drug or alcohol induced hypertension” section contains a dynamic list of any drugs the patient is currently taking that can contribute to secondary hypertension.^28,29^ In the aldosteronism section, the text will indicate whether the patient is on a medication that must be discontinued prior to screening or whether they are on a medication that does not need to be stopped but may impact screening results. The order for an ARR was automatically selected in patients with “increased” or “greatly increased” risk score.

### 2.4. Study design

The study evaluated whether a predictive model identifying a patient at risk of having an abnormal ARR result communicated via an alert and order set could alter ordering practices. Four departments that had physicians who were in informatics leadership were part of a pilot study, and these departments were excluded from the main study. All other adult primary care departments, those departments in the health system primarily providing primary care services staffed by family physicians, internists, nurse practitioners, and/or physician assistants, were randomized into a model group and a no model group. There were 77 departments in the model group and 76 in the no model group. These departments were in North Carolina and proximate regions of surrounding states. Since those patients with a risk score that was in the “greatly increased risk” range could have the alert triggered based solely on the risk model, patients were further divided based on whether they had a “greatly increased risk” score or not since this is expected to change the characteristics of the patients in the greatly increased risk model inclusion group.

Providers in both groups were identically informed of the availability of the secondary hypertension alert and order set via a weekly email that summarizes updates and changes to the electronic health record system. Beyond the initial email, there was no other proactive outreach to providers in either group. Data was collected for a period of eight months from September 2023 through April 2024.

### 2.5. Data collection

Each display of the alert and actions taken on the alert, including orders placed for secondary hypertension screening through the associated order set, were recorded in the electronic health record (EpicCare Ambulatory, Epic Systems, Verona, WI). Data extraction was performed with approval from the institutional review board of Novant Health which granted a waiver of informed consent given the nature of the study (22-2214).

### 2.5 Data analysis

The data was exported to Databricks for analysis using Python, version 3.13 (Python Software Foundation). The data was then summarized at a patient level, recording all actions taken on the alert across all times the alert was displayed. This summarization was necessary as the alert could be displayed multiple times within a single clinic encounter or across several encounters.

Information was summarized at the patient level in several steps. First, all entries associated after a patient’s first change were removed. This would either be a patient who switched from the no model to model clinic (or vice versa) or switched from the greatly increased risk to normal risk (or vice versa). Then for any patients who had an ARR ordered at some point (in their remaining entries), the first order was retained. Patients who did not have an ARR ordered retained the last entry before the above change occurred. The same process was repeated for the opening of the order set.

The percent of patients where the target actions, the launching of the order set and the ordering of an ARR, were taken was computed for both patients in the model and no model condition and was computed separately for patients in the highest risk group given that this group has a different logic for triggering the alert in the model group. Chi-squared tests were performed to determine whether the completion of the target actions was dependent on the model.

The patients in the model and no model departments were then compared to determine whether there were differences in sex, race and ethnicity, age, provider type, and department specialty using Chisquared tests. Finally, propensity score matching^30,31^ was performed to determine whether any differences between patients and providers in the model and no model groups would explain the differences in target action taken. Propensity score matching using K-Nearest Neighbors without replacement balanced the dataset for the relevant covariates listed below and then matched patients across the model and no model groups based on a patient’s probability of being in the treatment group.^32^ Statistical tests were conducted on both the unmatched and matched datasets with α=0.05 and effect size ϕ (0.1 is considered small, 0.3 medium, and 0.5 large).^33^

## 3. Results

### 3.1. Order set launches and ARR orders

Among patients who had predictive risk scores not in the top 1 percentile, 20493 patients had alerts displayed in the model clinics, the while 17820 patients had alerts displayed in the no model clinics. Of these patients, 438 (2.14%) in model clinics had the order set launched while 273 (1.53%) in no model clinics had the order set launched (P<0.001, ϕ=0.02). A total of 124 (0.61%) of these patients in model clinics had an ARR ordered and 34 (0.19%) patients in no model clinics had an ARR ordered (P < 0.001, ϕ=0.03).

Among patients who had predictive risk scores in the top 1 percentile, 2896 patients had alerts displayed in clinics where the risk score was displayed and used in triggering the alerts, the model clinics, while 1210 patients had alerts displayed in clinics where the risk score was neither displayed nor used for triggering alerts, the no model clinics. Of these patients, 63 (2.18%) in model clinics had the order set launched while 12 (0.99%) in no model clinics had the order set launched (P = 0.014, ϕ=0.04). A total of 19 (0.66%) of these highest risk patients in model clinics had an ARR ordered and zero patients in no model clinics had an ARR ordered (P=0.010, ϕ=0.04). Out of these 19 orders, 11 (57.89%) have results available with 3/11 (27.27%) of them returning an ARR > 35 while no results are available for the no model clinics. (Figures 3 & 4)

**Figure 3.**
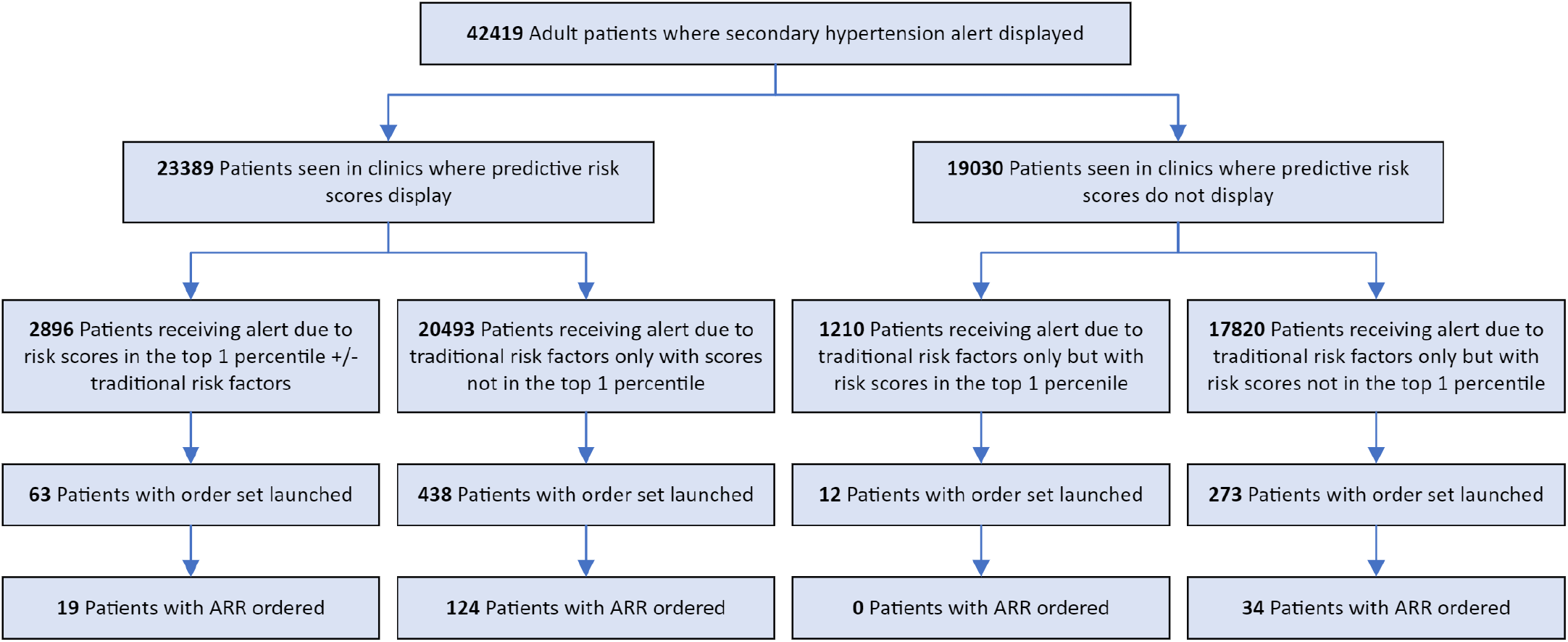
Study population with order set launch rates and ARR ordering rates.

**Figure 4.**
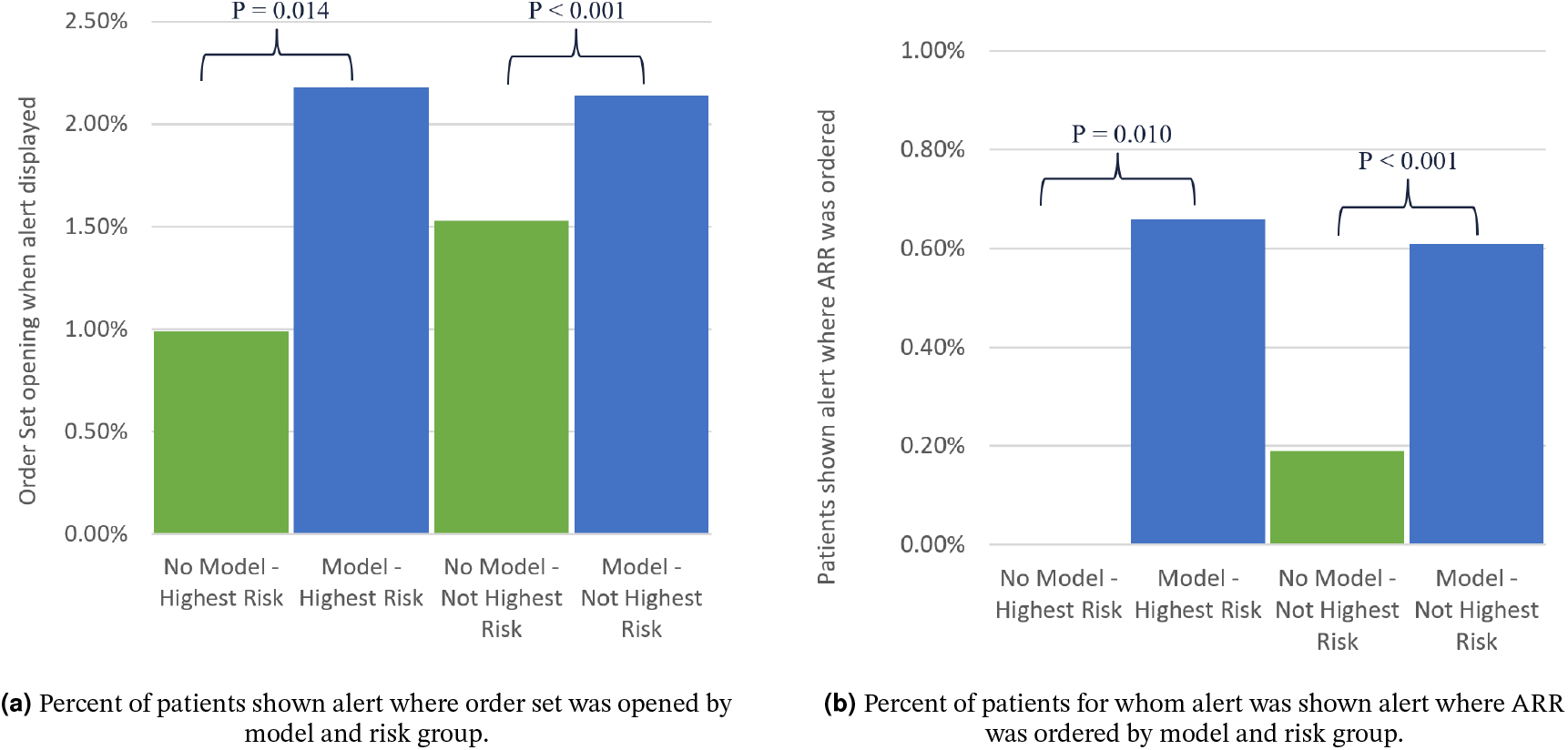
Impact of predictive model on order set opening and ARR ordering.

### 3.2. Group similarity

The patients included in the model and no model group were compared on the basis of sex, race and ethnicity, specialty of the visit department, visit provider type, and the calculated risk score at the given encounter, and independence was tested using the Chi-squared test. Additionally, the ages and risk scores represented in each group were compared using a T-test. There were no significant differences in sex representation between groups, but there were statistically significant differences in all other measures (Table 1).

**Table 1.**
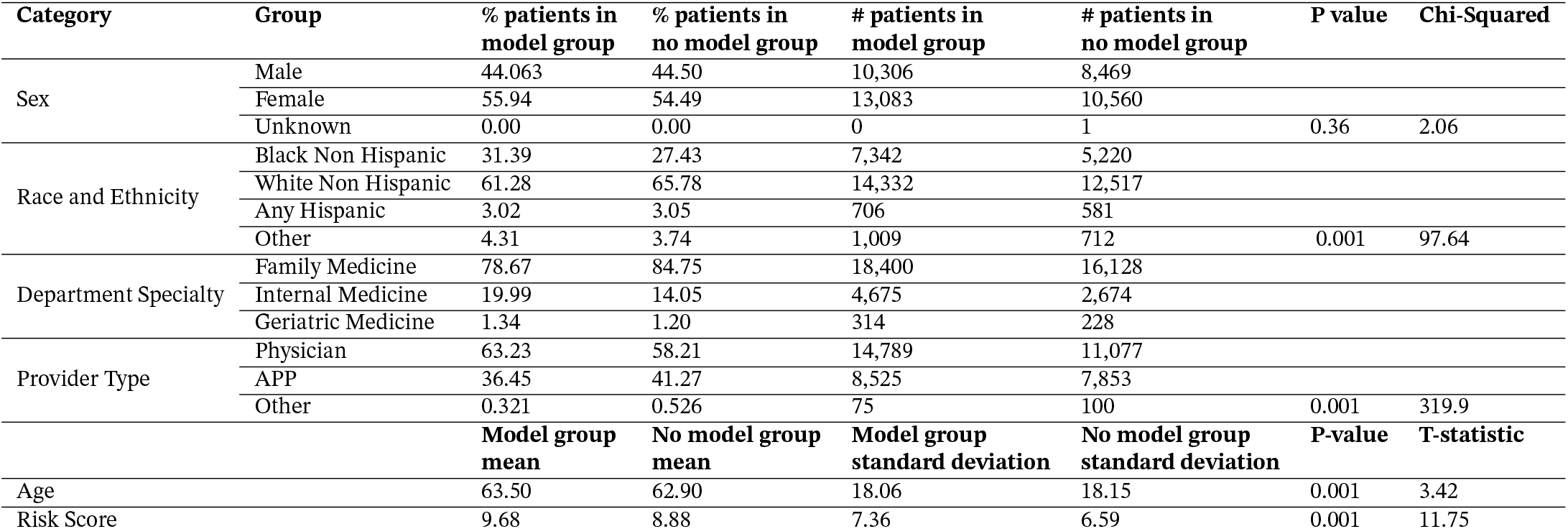
Differences in patient composition between model and no model group

When propensity score matching was performed to account for the differences between the model and no model group, a statistically significant difference was maintained for the difference in both order set usage (P<0.001) and ARR ordering (P<0.001) between the model and no model group for in the patients with risk scores not in the top 1 percentile. A statistically significant difference between the model and no model group was maintained for ARR ordering (P<0.001) and was established for order set usage (P<0.001) for patients with risk scores in the top 1 percentile. Quality of the propensity matching can be seen in figures 5 and 6.

**Figure 5.**
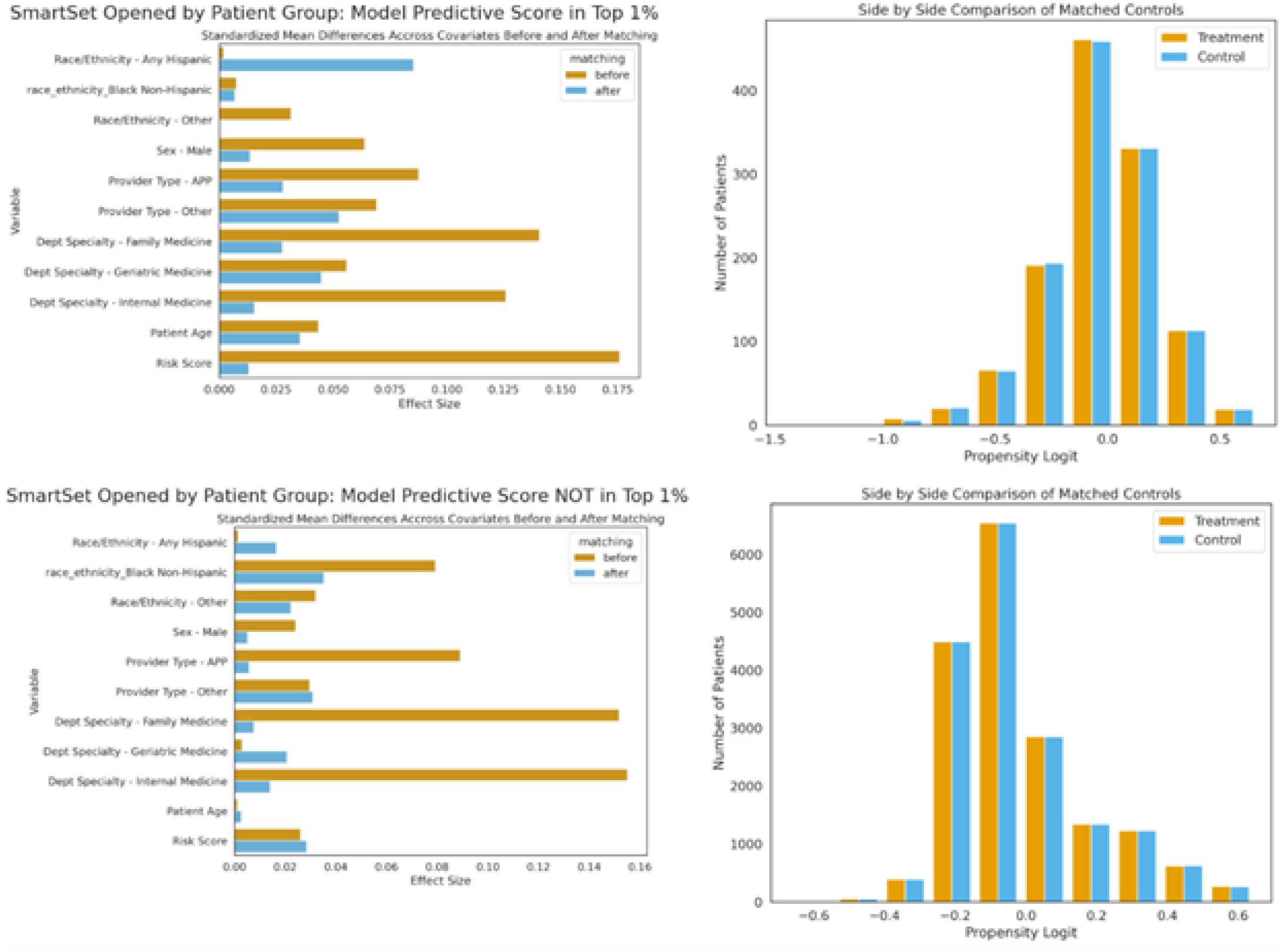
Propensity score matching results (order set).

**Figure 6.**
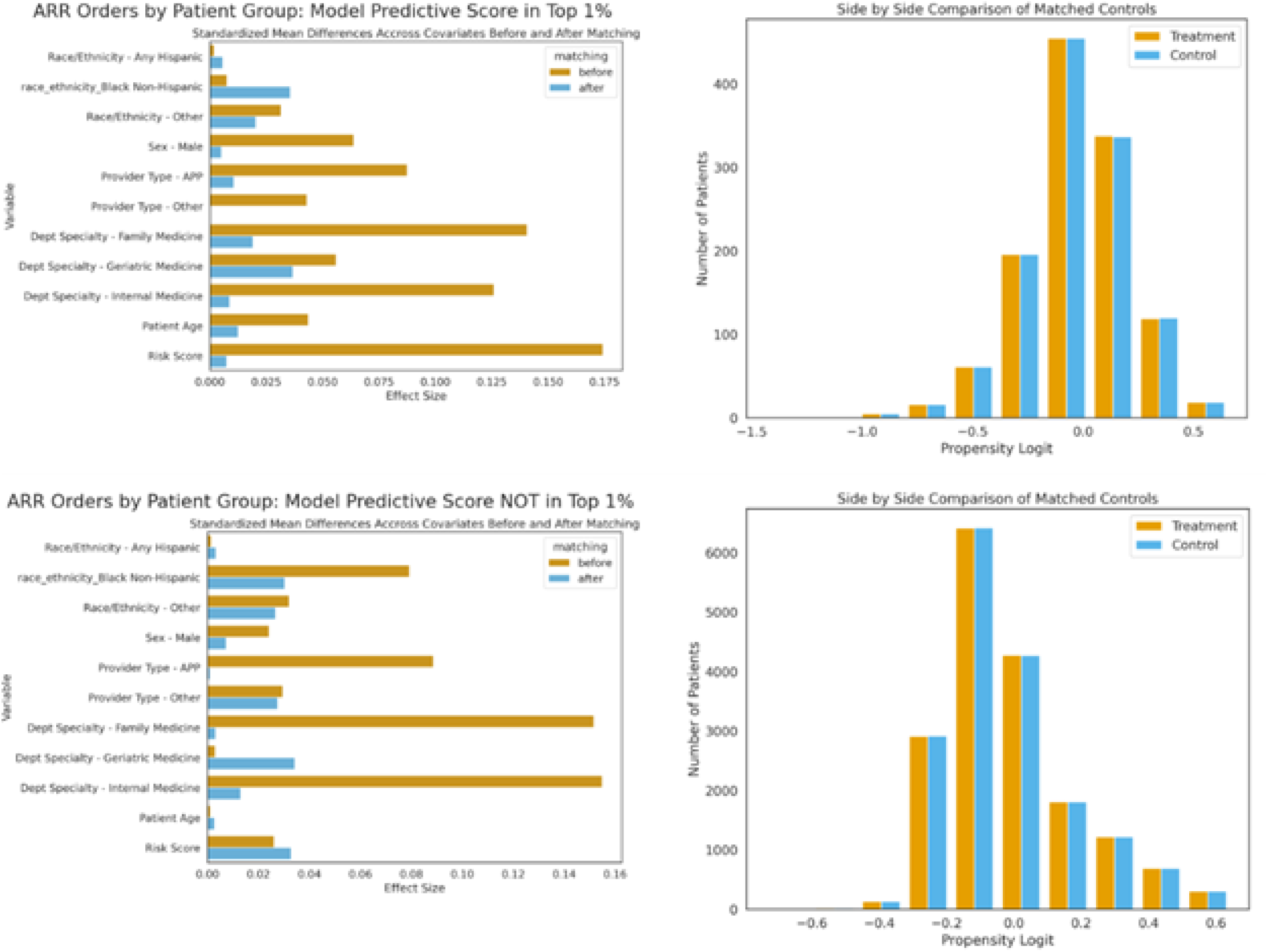
Propensity score matching results (ARR ordering).

## 4. Discussion

The goal of this study was to explore whether the addition of a predictive model to a clinical decision support system could help increase PA screening within the context of an already robust clinical decision support system. More generally, the integration of predictive models into clinical decision support systems is an important use case for a predictive model if the inclusion of the predictive model makes a meaningful impact on the desired use of the system.

Recommended PA screening is frequently missed in both the primary care^6,34-36^ and specialty care setting.^37^ While screening for PA lags behind many other recommended screenings, this is not likely to be out of malice or carelessness, but rather due to the variety of clinical findings that can suggest primary aldosteronism being stored in disparate areas of the chart along with the complexity of incorporating secondary hypertension screening guidelines into an already busy office visit. Ascertaining the risk of secondary hypertension requires elements from the vital signs, lab values, social history, and medication list which are generally stored in multiple parts of the health record.

The fundamental theorem of biomedical informatics states that “A person working in partnership with an information resource is ‘better’ than that same person unassisted”.^38^ Previously described clinical decision support tools support using clinical decision support tools to increase screening for PA, but there is still significant room for improvement in increasing PA screening rates.^13^ This work supports the addition of a predictive model to the clinical decision support system to further increase the rate of PA screening. The work of gathering data from multiple parts of the chart and forming a risk estimation can be performed by the risk model, thus decreasing the cognitive burden of the clinician. A potential adverse impact of any call to improve screening for any condition is the increased cognitive burden associated with the decision to screen and the act of screening. Increased cognitive burden is associated with burnout in physicians^39^ and medical errors in nursing.^40^

The addition of risk models to the electronic health record has the potential to further increase the cognitive burden of healthcare providers. This work demonstrates that incorporating the risk model into existing clinical decision support tools may increase the uptake of those tools but does not explicitly address the cognitive burden of the model and the CDS tools. Using predictive analytic models to automatically select further screening tests within a CDS tool limits the clinician’s decision making to identifying times where the model may not accurately represent the patient’s risk rather than requiring the clinician to read the results of the predictive model and independently order appropriate additional testing.

### 4.1. Limitations

There are multiple etiologies of secondary hypertension, and this study only looks at PA. While the efficacy of a predictive model in increasing PA was demonstrated, the efficacy of predictive models for screening for other etiologies of secondary hypertension was not evaluated. A comprehensive suite of risk models for multiple causes of secondary hypertension incorporated into a secondary hypertension clinical decision support tool has the potential to provide additional value for identifying multiple etiologies of secondary hypertension, but this has not been clearly identified by this study.

The path from identification of possible PA to confirmation of PA to lateralization of aldosterone secretion to definitive management with surgery^5^ will take several months at minimum and may take a year or more when applied practically, particularly when scheduling of elective adrenalectomy is limited based on both patient availability for an elective surgery and the availability of subspecialty surgeons to perform adrenalectomies. For these reasons, this study did not examine the impact of adding a risk model to clinical decision support systems on long-term blood pressure control as the impact on longterm blood pressure control may not be realized for a year or more.

Additionally, this intervention is expected to make a significant impact on a relatively small fraction of patients with hypertension, but since most patients with hypertension do not have PA, the overall impact on hypertension control in the study population may be minimal.

## 5. Conclusions

The addition of a predictive model to a PA clinical decision support tool significantly increases the rate of screening for PA, an underscreened condition. With the current proliferation of predictive models for PA, operationalizing these models through decision support tools should be a key priority. The integration of predictive models with clinical decision support tools may help to increase screening for other under-screened conditions. Future studies should examine the generalizability of this approach to a wider range of under-screened conditions.

## Data Availability

Patient level data used in the study is not available as a waiver of informed consent was given by the institutional review board given the nature of clinical decision support studies.

## 6. Author contributions

**Peter Mack:** Conceptualization, Methodology, Software, Investigation, Writing – original draft. **Casey Cole:** Methodology, Software, Formal analysis, Data Curation, Writing – Review & Editing. **Mintaek Lee:** Methodology, Software, Formal analysis, Data Curation, Writing – Review & Editing. **Lisa Peterson:** Conceptualization, Project administration, Writing – Review & Editing. **Matthew Lundy:** Conceptualization, Supervision, Writing – Review & Editing. **Karen Hegarty:** Conceptualization, Supervision, Writing – Review & Editing. **William Espinoza:** Conceptualization, Methodology, Software, Formal analysis, Investigation, Data Curation, Writing – Review & Editing.

Additional support with formal analysis and data curation provided by **Vinay Palagiri** and **Yuryi Malakhau**.

